# Factors influencing the provision of antenatal urine testing in Kavrepalanchowk, Nepal: a mixed-method study

**DOI:** 10.1101/2025.09.03.25335067

**Authors:** Rajani Shakya, Abha Shrestha, Samita KC, Prasanna Rai, Chanda Thakur, Abha Shrestha, Emma Radovich

## Abstract

Routine urine testing during pregnancy is important for diagnosing complications like pre-eclampsia and gestational diabetes, but it is among the least frequently performed antenatal care (ANC) components in settings like Nepal. This study explores the infrastructure, supply, training, and other obstacles to routine urine testing in ANC in Nepal.

Our mixed-method study analyzed the nationally representative Nepal Health Facility Survey (NHFS) 2021 and primary qualitative data from Kavrepalanchok district, Nepal: in-depth interviews (n=18) with local policymakers, ANC providers, and pregnant women and semi-structured observations in six health facilities. Facility observation notes and interview transcripts were coded using a template analysis approach, and the NHFS 2021 was descriptively analyzed. Findings were mapped to the WHO framework for the quality of ANC.

Among health facilities offering ANC services in the NHFS 2021, only 32.5% performed urine dipstick testing on-site. Physical infrastructure and supplies enable pregnant women to provide urine samples and was adequate at health facilities. However, ANC providers rarely performed point-of-care dipstick tests in lower-level facilities. Limited laboratory capacity in lower-level facilities resulted in pregnant women making repeated visits or being referred to higher-level facilities to obtain urine tests. Healthcare providers were aware of the significance of urine testing but not of the recommended frequency. Urine tests were regularly performed during the first ANC visit and repeated only if there were other symptoms of pregnancy complications.

Despite the availability of adequate physical resources for urine testing, routine urine tests were not conducted in every ANC visit, as recommended in Nepal guidelines. Training and supportive supervision are necessary to modify urine testing practices in ANC, including performing dipstick tests by ANC providers, at the point of care, in rural health facilities lacking lab services.

## Introduction

Urine testing is regarded as an integral component of antenatal care (ANC) services(1). Routine urine testing in pregnancy is an effective screening tool that is done to detect levels of albumin, nitrates, and glucose in the urine, helping to identify pregnancy complications such as gestational diabetes mellitus, preeclampsia, asymptomatic bacteriuria, and other infections as well(2–4). The 2016 WHO antenatal care model recommends urine analysis to be conducted during each ANC visit(5). In 2022, ANC to PNC continuum of care guideline was published by Nepal’s Family Welfare Division indicating repetition of urine tests in every ANC visit for proteinuria(6). A study examining coverage and content of ANC in low-income and middle-income countries found blood pressure measurement was the most commonly reported component, and urine test and information on complications were the least(7). Similar finding was there from a study in rural Bangladesh, where urine and blood tests are done less often than other ANC checks like blood pressure and weight (8).

Commercially available dipsticks have evolved to be a highly efficient tool for investigating, detecting, and screening diseases with rapid, high-quality results, maintaining ease of use and have been used in clinics & homes without the help of central laboratory devices or advanced expertise(4,9,10).

Evidence suggests that inadequate water, sanitation, and hygiene (WASH) adversely affects maternal and reproductive health, as well as fetal and newborn outcomes (11). Unfortunately, in many low-income settings, many women receive maternity care in facilities without sufficient WASH capabilities, especially with the less availability of water and soap (12–14). While WASH in facilities is increasingly recognized for delivery care, few appraisals consider how WASH impacts antenatal services(15,16). The Nepal National Standard for WASH in Health Care Facilities has documented guidelines for WASH infrastructure, including standards for toilets, washbasins, running water, soap, and proper waste disposal which are necessary for providing quality ANC services and ensuring the health and safety of pregnant women (17). However, the 2021 Nepal Health Facility Survey found variation in the performance of urine testing during ANC at the different health facility levels (18). The major factors driving inconsistencies in the performance of antenatal urine testing in Nepal has not been well examined. This study aimed to explore the infrastructure, supply, training, and other obstacles to routine urine testing in ANC in Nepal.

## Methods

### Ethical Statement

Ethical approval for the primary data collection was obtained from the Nepal Health Research Council (Ref no. 984), the Institutional Review Committee of Kathmandu University (Ref no. 85/24), and the research ethics committee of the London School of Hygiene and Tropical Medicine (29823).

Ethical approval was not required for the secondary analysis of NHFS 2021 data, which were obtained with permission from the Demographic and Health Survey Program website (www.dhsprogram.com). In the original survey, ethical clearance was obtained from the Ethical Review Board of Nepal Health Research Council and ICF International(18). Written informed consent was obtained from the facility head for health facility observations and from the eligible health workers and clients of selected health facilities.Verbal consent was taken from pregnant women and health care providers to observe ANC consultation.

### Conceptual Framework

We organized our investigation into antenatal urine testing around the WHO framework for the quality of ANC(19)(Fig1). This framework is intended to assist healthcare providers, managers, and policy-makers to better understand and assess health system characteristics required to deliver and improve the quality of routine ANC for a positive pregnancy experience(20). We adapted the WHO framework for the quality of ANC to focus on urine testing during ANC visits, with particular attention to: provision of care, experience of care, essential physical resources, and competent, motivated human resources(19). Data included in the analysis were primary data collection of in-depth interviews and health facility observations and a secondary data analysis of the 2021 Nepal Health Facility Survey.

**S1 Fig1:**
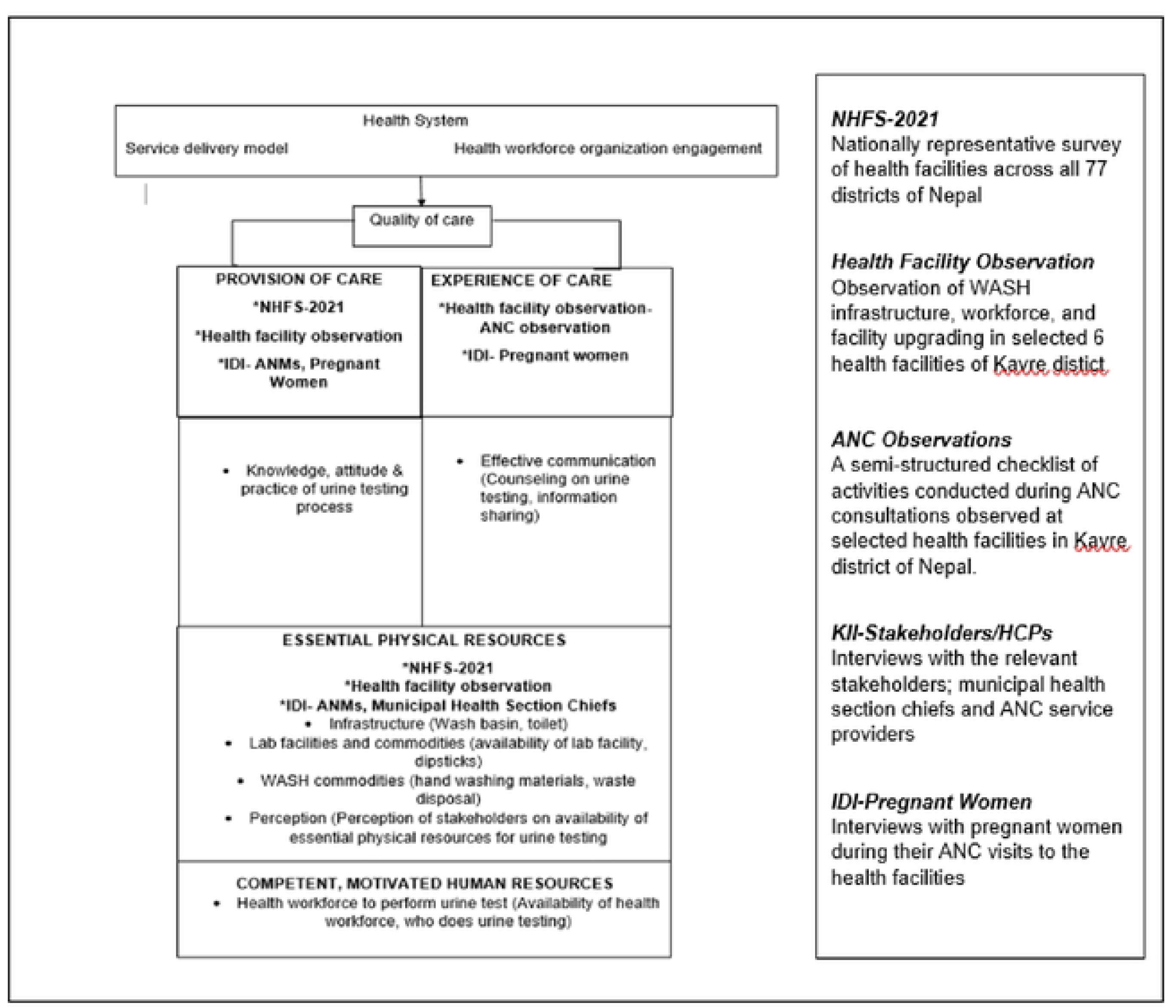
Data Sources included in the analysis mapped to the WHO framework for the quality of ANC in terms of urine testing. ANC =Antenatal Care, KII-Key Informant Interview, IDI-In Depth Interview

### Study design

This convergent, mixed-method study drew data from in-depth interviews with relevant stakeholders (municipal health section chiefs, healthcare providers, and pregnant women) and observations of health facility infrastructure and ANC consultations in the Kavrepalanchok district, and a secondary analysis of the nationally representative Nepal Health Facility Survey (NHFS) 2021(18).

### Data Collection

#### Primary data

Primary qualitative data were collected from three rural and urban municipalities in Kavrepalanchok district. We purposively selected the municipalities, and the governmental health facilities within the municipalities, in conversation with health section chiefs to include a range of types of primary-level health facilities, including those with birthing facilities and laboratory services, those with higher ANC client load, and those with plans to or that had recently been upgraded as part of efforts to build more municipal hospitals (21, 22). The study sites were the different levels of health facilities: municipal hospital, Primary Health Care Centers and Health Posts situated in these municipalities.

Qualitative data collection was done by two researchers (RS and SK) between February-June 2024 and included facility-based data collection (observations and interviews) and interviews with municipal officials. Facility-based data collection included non-participatory health facility and ANC consultation observations, and interviews with auxiliary nurse midwives (ANMs) and pregnant women. Researchers visited each facility for 2-3 days to observe urine testing capabilities of health facilities and practices during ANC visits using a semi-structured checklist (S2 Facility Observation Tool). The semi-structured observation questions were adapted from the WHO monitoring framework for delivery car, alongside ethnographic approaches of participant observation for documenting the process of ANC provision and urine testing (23,24).

Interviews were conducted face-to-face with ANMs providing ANC services and pregnant women receiving the ANC services during facility-based data collection, and with municipal health section chiefs in their offices. Separate interview guides were developed for each type of interview participant. Pretesting of interview guides for pregnant women and health care providers was done in one of the tertiary-level hospitals in the Kavrepalanchok district and adapted accordingly. The semi-structured observation checklist was also pre-tested in the same tertiary-level hospital and adapted accordingly. The research team iteratively adapted the interview guides based on emerging findings and reflection notes during data collection.

#### Secondary data

The NHFS 2021, a nationally representative cross-sectional survey, collected data from health facilities across all 77 districts of Nepal. These included government-managed facilities (Federal and Provincial level hospitals, local level hospitals, Primary health care centers (PHCCs), Health posts (HPs), Urban Health centers (UHCs), community health units (CHUs), and HIV testing and counseling clinics, as well as private not-for-profit organizations, non-governmental organizations, for-profit organizations, and mission/faith organizations. The survey used a stratified random sample of 1,633 health facilities from a pool of 5,681 eligible facilities, selected through equal probability systematic sampling with sample allocation. Facilities were grouped by type within each province for stratification(18). The sample allocation aimed for consistent survey precision across provinces. For this analysis, we included 1,538 health facilities that offered ANC services. NHFS data collection occurred between January 27, 2021, and September 28, 2021, with a pause from May to July due to COVID-19 lockdowns.

We used data from the ‘Facility Inventory Questionnaire’ and ‘Health Provider Questionnaire’ from NHFS 2021(18). Among all facilities offering ANC services, we examined the service availability, competent human resources and essential physical resources for providing urine testing in pregnancy (Table 1). Service availability referred to whether urine testing was offered at the facility via dipstick tests (or whether urine samples were sent outside the facility) and the specific routine urine tests offered in ANC. The assessment of service availability included facilities that reported generally offering a specific routine urine test in ANC even if the facility lacked a test kit in stock on the day of the survey (Table 1). Competent human resources included whether at least one ANC provider at the facility had received any ANC service training or training in ANC screening within the past 24 months. Essential physical resources included WASH infrastructure (availability and condition of client toilets, availability of hand washing materials, and waste disposal systems) to collect and handle urine samples and whether a valid test kit was available among facilities that reported performing urine testing on site and that reported offering the corresponding test in ANC.

**Table 1.**
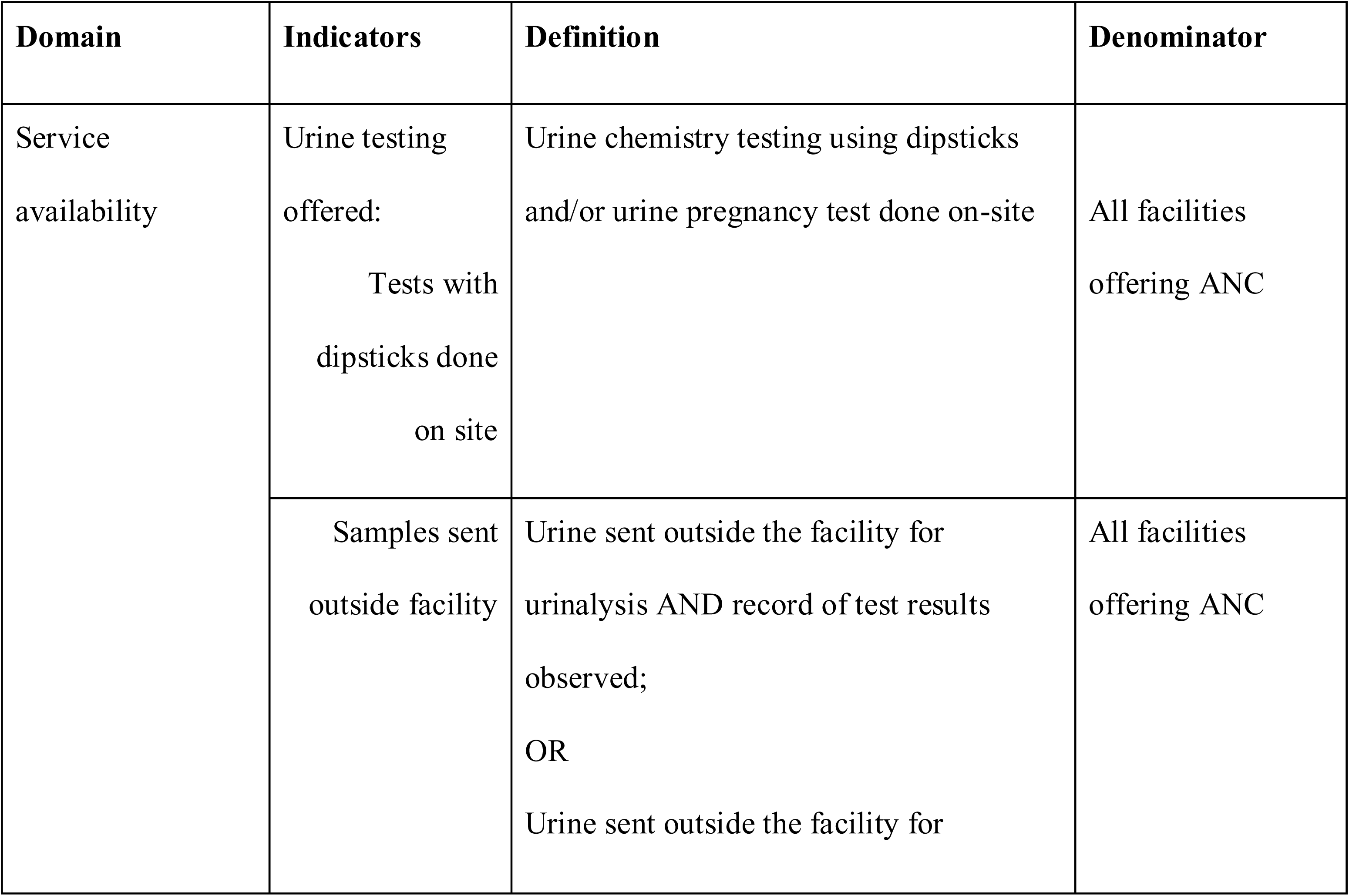
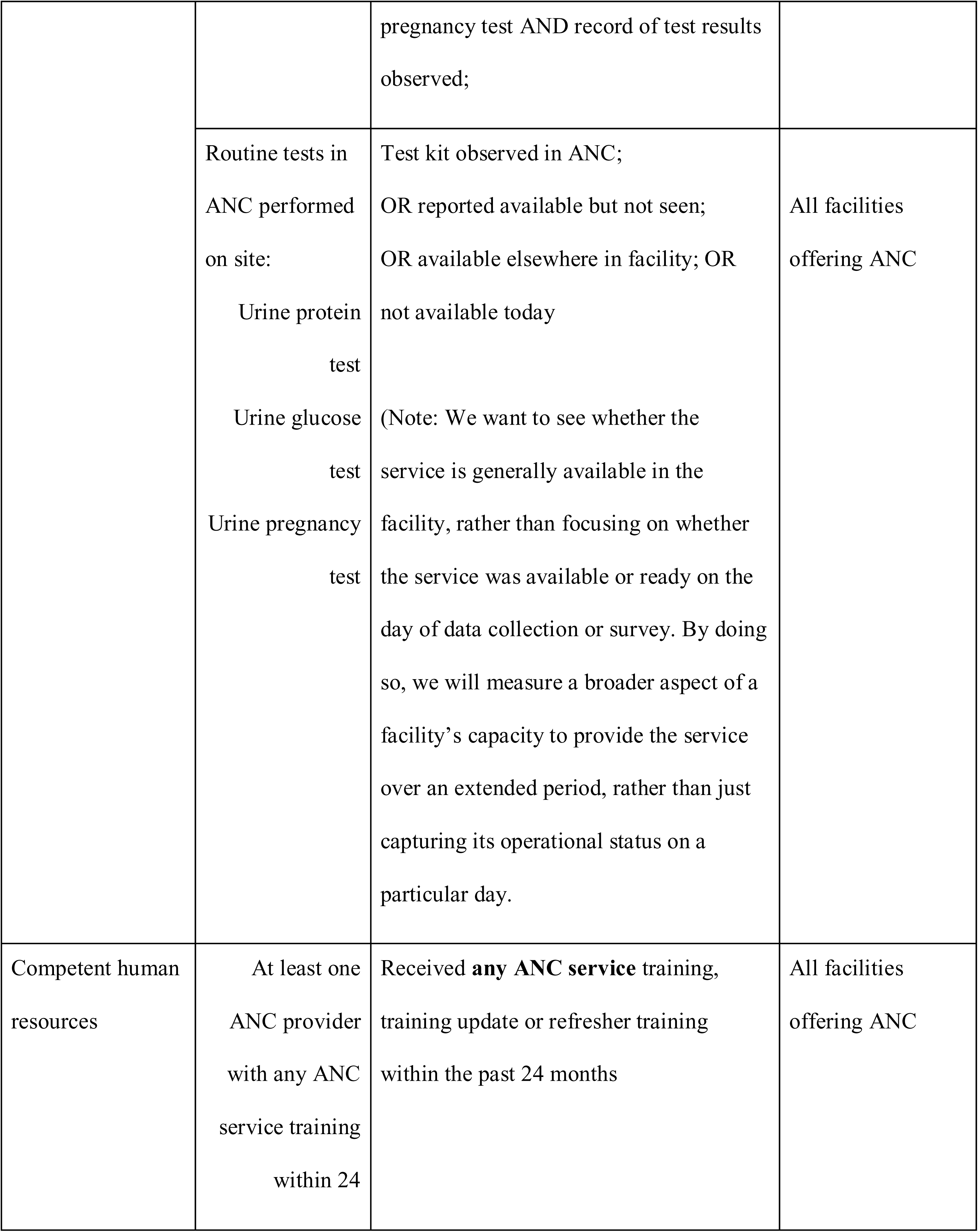
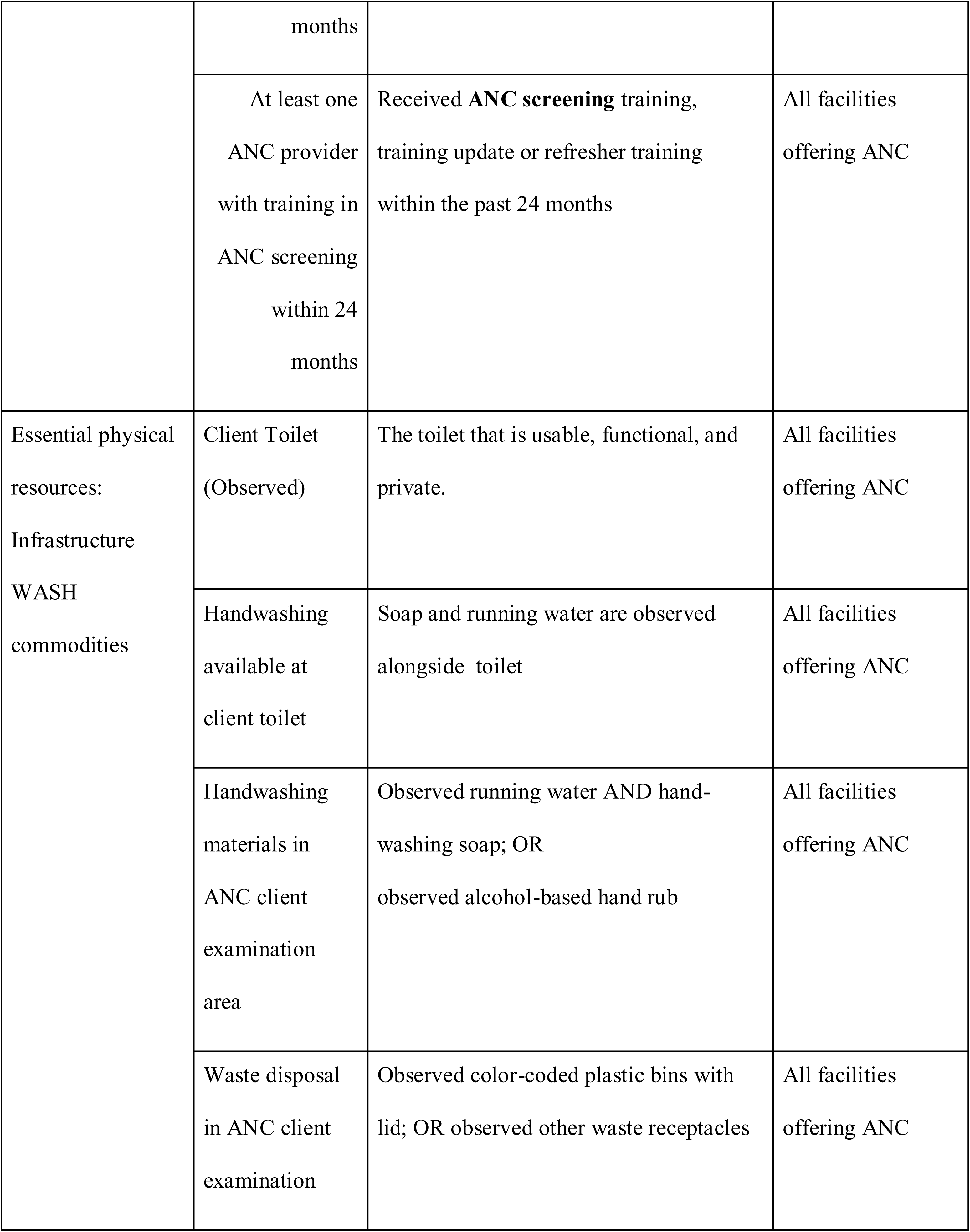
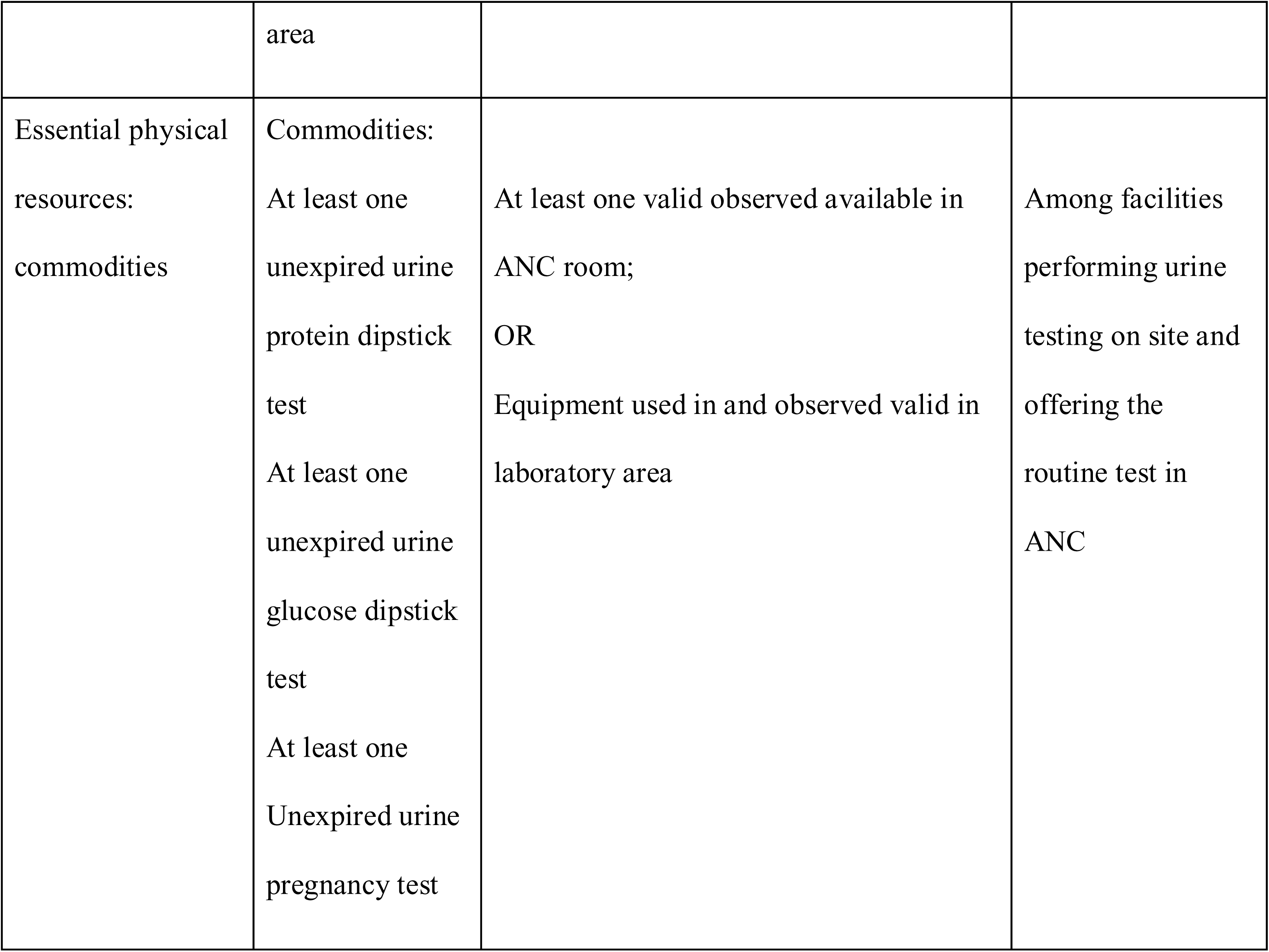
Operational definitions of the indicators of the study from Nepal Health Facility Survey 2021.

### Data analysis

Recordings from the interviews were transcribed verbatim in Nepali. Qualitative interview transcripts and notes from the facility and ANC observations were analyzed using template analysis. Template Analysis is a form of thematic analysis that tends to define themes and initial coding structure from a sub-set of data (24). Therefore, the codes came out from our *a prior* interests in using the WHO framework for the quality of ANC, alongside an initial reading of the data. The iterative analysis process involved initial coding, coding consensus and refining codes and themes through weekly meetings of the research team to discuss emerging findings. Four members of the research team (RS, AS, AS, and ER) had previously worked together on an antenatal quality improvement implementation study, also in Bagmati Province; the researchers drew on this experience to reflect on findings and considered how our perspective and previous knowledge may have impacted interpretation in this study(25). Relevant excerpts from the transcripts were translated into English for the write-up of findings.

We conducted a descriptive analysis of the NHFS data to calculate the percentage of facility types meeting the indicators for urine screening in pregnancy. All analyses were weighted to account for the complex, clustered sample design of the 2021 NHFS. We present the unweighted sample size of facilities by type and by province to show the actual number of health facilities contributing data to the analysis. Data analysis was conducted using STATA 14 (StataCorp, Inc).

We triangulated findings from the qualitative and quantitative data, drawing on the framework for quality ANC (Figure 1). Themes were considered important where there was consistency across the primary and secondary data, with discrepancies interrogated and discussed. To enhance the validity and credibility of our study, we presented initial findings with key stakeholders and research participants at a workshop in July 2024. Reflections from workshop participants contributed to refining themes and the interpretation of results.

## Results

We brief describe the health facilities and interview participants contributing to the study, and then present integrated findings from the primary and secondary data analysis under four components of the framework: provision of care, experience of care, essential physical resources and competent, motivated human resources (Figure 1).

Primary data collection was conducted in six health facilities across the three selected municipalities and interviews were conducted with 18 participants (Table 2).

**Table 2:** Study setting and number of study participants for the primary qualitative data collection in three municipalities in Kavrepalanchok District.

| Municipalities | No. & Types of Health Facilities Observed | Interviews with municipal health official | Interviews with Pregnant women | Interviews with Health Care Providers |
| --- | --- | --- | --- | --- |
| Municipality A(Urban) | Municipal Hospital(n=1), Health Post(n=1) | 1 | 2 | 3 |
| Municipality B(Urban) | Primary Health Care Center(n=1), Health Posts(n=1) | 1 | 4 | 2 |
| Municipality<br>C(Rural) | Health Post(n=2) | 1 | 2 | 2 |
| **Total** | **6** | **3** | **8** | **7** |

Table 3 compares six health facilities (HF1 to HF6) on key infrastructure and practice indicators, based on the facility observations and interviews with healthcare providers. All the facilities observed were birthing units except for one (Table 3).

**Table 3:** Provision of care, physical resources and human resources for antenatal urine testing among the six facilities observed for primary data collection.

| **Description/ Health Facilities** | **HF1** | **HF2** | **HF3** | **HF4** | **HF5** | **HF6** |
| --- | --- | --- | --- | --- | --- | --- |
| **Provision of Care** |  |  |  |  |  |  |
| No. of ANC Providers | 3 | 1 | 3 | 2 | 2 | 2 |
| Birthing Unit | Yes | No | Yes | Yes | Yes | Yes |
| Urine testing practice | Once in each trimester | Not Performed | First ANC visit, then as per need | First ANC visit, then as per need | Each ANC visit | First ANC visit, then as per need |

| **Essential Physical Resources** |  |  |  |  |  |  |
| --- | --- | --- | --- | --- | --- | --- |
| At least one functional toilet and handwash basin | Yes | Yes | Yes | Yes | Yes | Yes |
| Lab | Yes | No | Yes | Yes | Yes | Yes |
| Availability of Urine dipstick strips | Yes | No | Yes | Yes | Yes | Yes |
| **Competent Human Resource** |  |  |  |  |  |  |
| Availability of lab personnel at the point of observation | Yes | No | Yes | Yes | No | Yes |

In the 2021 NHFS, 1538 health facilities reported offering ANC services and were included in the assessment of antenatal urine testing (Table 4).

**Table 4:**
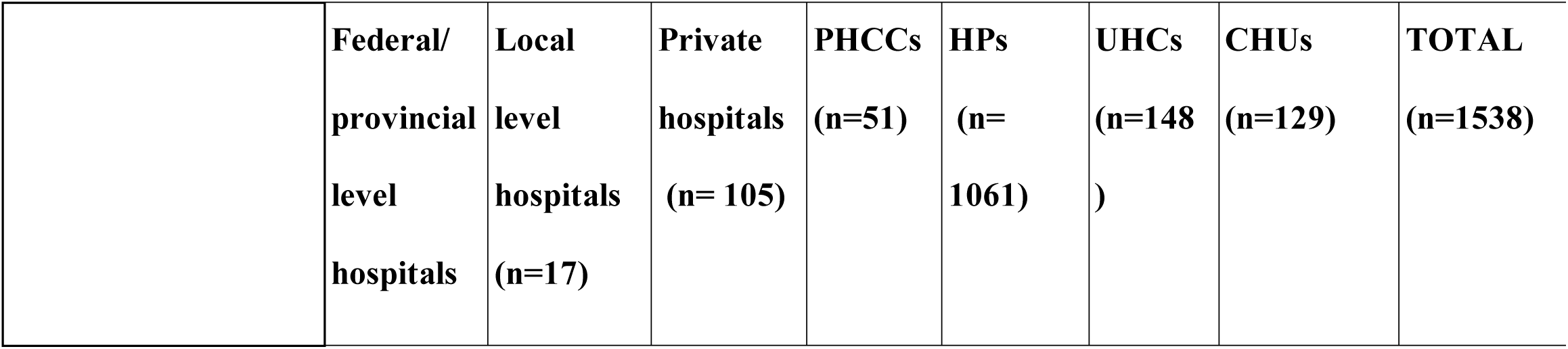
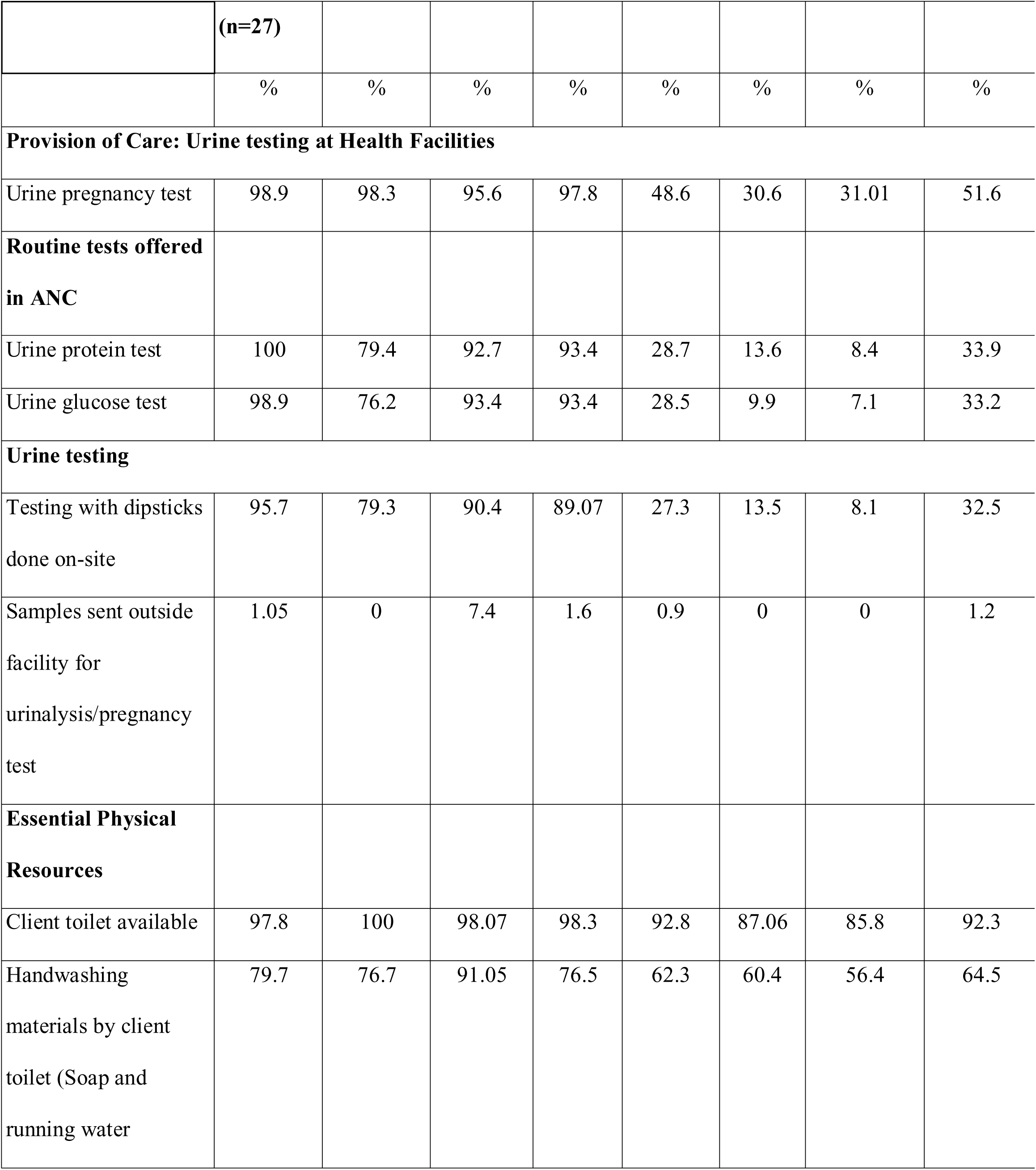
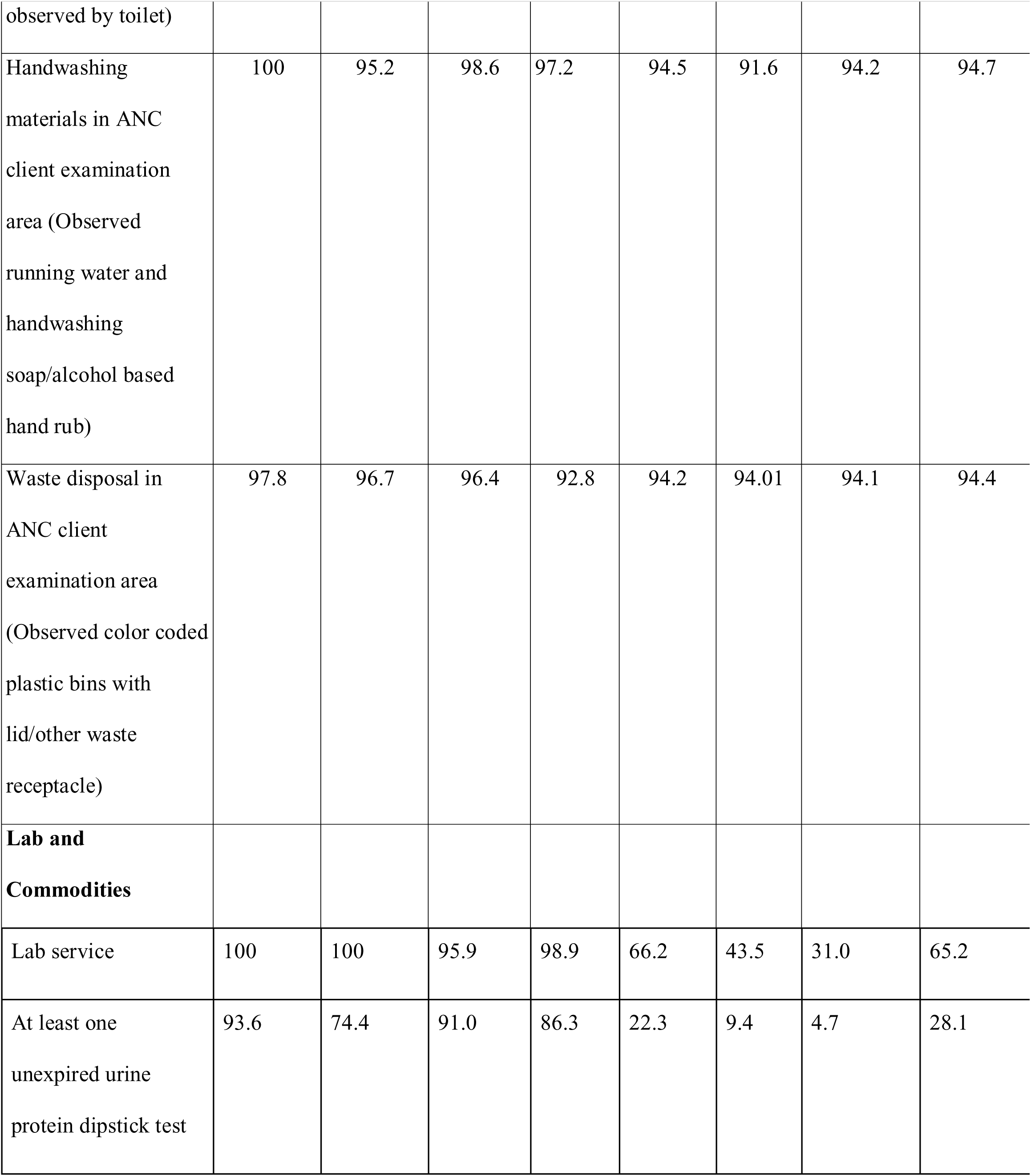
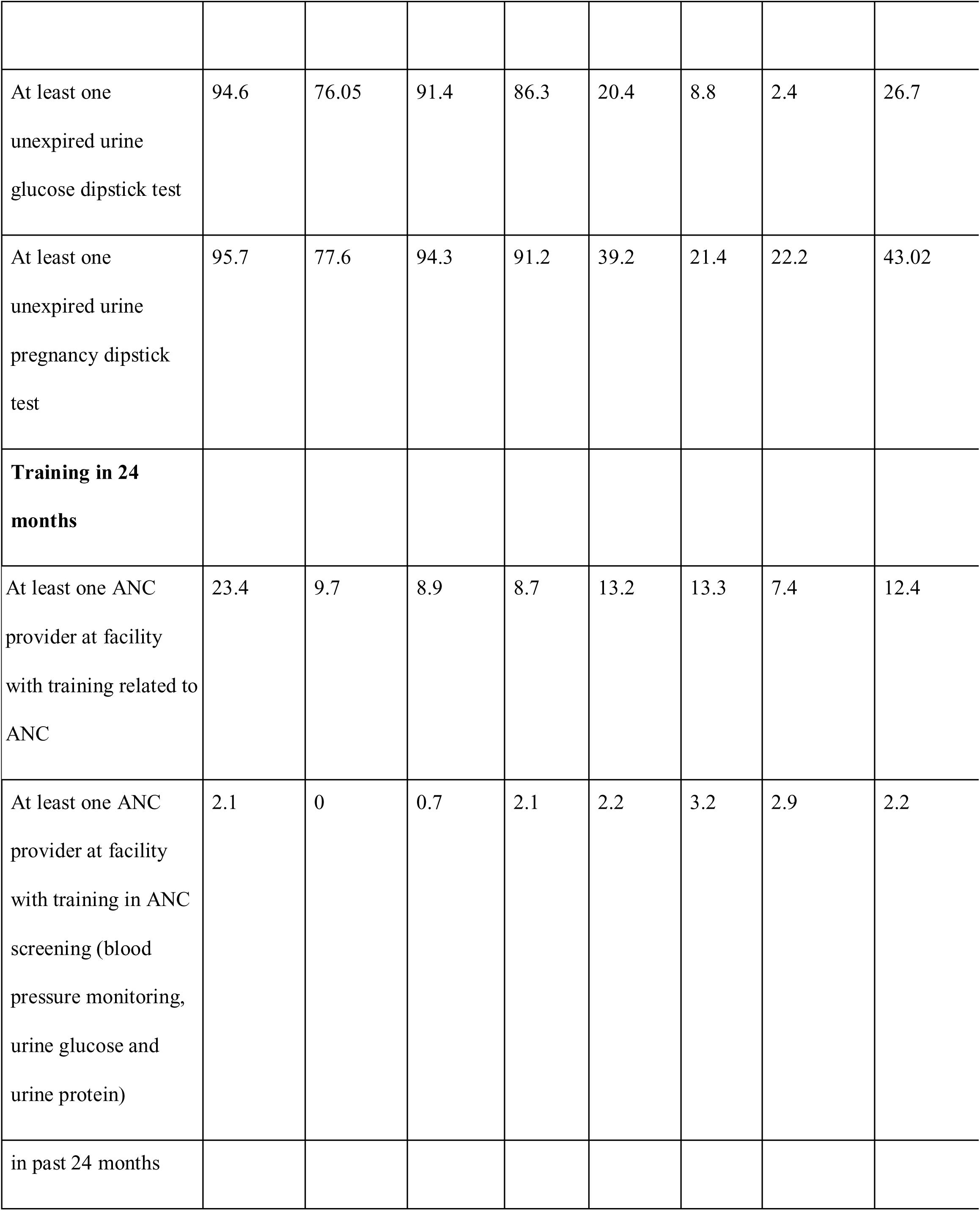
Provision of care, physical resources and human resources in health facilities offering ANC services in the 2021 Nepal Health Facility Survey (unweighted n=1538)

### Provision of Care: antenatal urine testing practices

Urine tests were generally performed by lab personnel, rather than ANC providers. Urine testing varied among the facilities observed in Kavrepalanchok, with three of six facilities reporting performing urine tests once during the first ANC visit and then as per need and one facility not offering any urine testing (Table 2). Only one of the health facilities was observed performing urine tests in each ANC visit in line with the guideline; in this facility, the ANM used dipstick tests at the point-of-care during ANC visits. Secondary data analysis of the NHFS showed that performance of dipstick tests done onsite for urine varied between facility levels, with 89.1% of PHCCs but only 27.3% of HPs performing urine dipstick testing onsite (Table 4).

Nearly all ANMs interviewed described performing a urine test for proteinuria and glycosuria at the 12-week pregnancy confirmation visit and only repeating urine tests if symptoms were reported.

> *“We usually do the urine test initially for pregnancy. If at that time, they mention that they have a burning sensation during urination or suspect a urinary tract infection, we will conduct the urine test then as well. Otherwise, we do it at 12 weeks when pregnant women come for the check-up. After that, we do it again at 24 weeks. If any infection is detected, we will test whenever necessary, as per your symptoms. Finally, we conduct the test between 36 and 38 weeks. So, in total, we do it two or three times.’’ (Healthcare provider-1)*

### Experience of Care: effective communication for counseling on urine testing

In the interviews, ANMs mentioned that they were providing counseling to pregnant women on urine testing. However, in-depth interviews with pregnant women depicted a lack of communication by ANMs about antenatal urine testing. Most of the pregnant women did not recall receiving guidance for the urine sample collection, nor the results of the test. On observation, researchers also did not see any ANMs offer guidance to pregnant women on giving a urine sample or explain the result.

> *“No, they didn’t mention anything at the lab either. After the checkup, they told me the report would be ready in two hours. I waited and received the report. When I brought it to them (the ANM), they (ANM) said it was okay and good, but when I asked what it was for, they didn’t provide any details. (Pregnant Women 2)*

### Essential physical resources

#### WASH Infrastructure and Commodities

At least one functional client toilet along with running water was available in all six health facilities observed, but soap or hand wash was not available alongside the client toilet at any of the health facilities (Table 3). Handwashing materials were available elsewhere in the facility, such as in The ANC room. Researchers also noted the presence of gender-separated toilets in municipal hospital along with commode-style toilets in recently constructed infrastructure, compared with a pan style in the older infrastructure. Pregnant women interviewed expressed satisfaction with the facilities.

> *It’s very clean here, and it feels like home as soon as you arrive. The toilet was fine, with water for handwashing and dustbins for disposal. (Pregnant Women 4)*

Secondary data analysis of the NHFS also showed more than 85% of all facilities had functional toilets for clients (Table 4). However, there was lower availability of soap and running water next to client toilets in some facilities, such as Health Posts (62.3%) and CHUs (56.4%), though hand-washing materials were available in the ANC client examination areas in nearly all facilities.

#### Lab facilities and commodities

Lab facilities were available at all health facilities observed, except for one health post (Table 3), where urine testing was generally done by ANMs. The availability of lab facilities varied by facility evel in the NHFS analysis, with nearly universal availability in government hospitals and PHCC, yet only 66.1% of health posts had a lab (Table 4)

Among the six facilities observed in Kavrepalanchok, urine dipstick test for proteinuria were reported available at the municipal hospital and PHCC; however, it was not available at one of the health posts in the urban municipality. Valid protein dipsticks and glucose dipsticks were observed in the ANC room or lab. In the national-level data, urine protein and urine glucose tests were offered in more than 90% of PHCCs, and 86.3% of PHCCs had unexpired urine protein and urine glucose tests in stock (Table 4). However, among health posts, urine protein and urine glucose tests were less often routinely offered in ANC, and only 22.3% and 20.4% of health posts had unexpired urine protein and urine glucose test kits, respectively, in stock (Table 4).

In the interviews, ANMs did not mention any issues with logistics for urine test supplies. However, one health section chief discussed barriers with an insufficient supply of dipsticks from the province and lack of budget to purchase at the municipality.

> *“ It seems like we don’t have enough dipsticks. We’re trying to purchase them, but just buying dipsticks isn’t enough—we need to buy everything. The budget is limited, very minimal. The district health office is supposed to supply us with these because the province government is responsible for purchasing the testing kits. However, they haven’t arrived on time, which has caused this issue.” (Policy maker 003)*

### Competent, motivated human resource

The qualitative investigation found ANM’s were aware of the importance of urine tests, including that proteinuria could indicate preeclampsia, but lacked knowledge about the guideline-recommended repetition of urine tests at each ANC visit. Analysis of NHFS found less than 4% of all facilities had at least one provider who had received any training on screening tests in ANC in the last 24 months (Table 4).

Despite the availability of lab services in five of the six facilities observed, lab personnel were present in only four health facilities during the observation (Table 3). There was also day-to-day variation in the availability of lab technicians at the facilities, with one of them observed remaining until 2pm, although the office hours are until 5pm (Health Facility Observation) and in one health post in a rural municipality, the lab technician was only scheduled to be on site once a week. Researchers observed ANMs performing urine dipstick tests in this health post, where there was also a handmade wall chart mentioning the urine test and other tests to be done at every ANC visit.

The unavailability of lab personnel and ANMs relying on lab technicians for point-of-care testing was also one of the barriers to providing regular urine testing services to pregnant women.

> *“When I was attending Family Planning Association at Banepa to do VIA(Visual inspection with acetic acid), I saw this protocol on wall, then I asked my junior to make it who are more knowledgeable and pasted in wall. That’s how we have been doing.”-Health Care Provider 006*

## Discussion

This study aimed to assess the factors shaping provision of urine testing during ANC in Nepal, focusing on provision of care, experience of care, essential physical resources, and competent, motivated human resources. Regarding provision of care, we found wide variation in the performance of urine dipstick testing by facility level and that urine testing was generally done by lab technicians. Regarding experience of care, we found little evidence of communication between healthcare providers and pregnant women around the process and purpose of urine testing during ANC. Regarding essential physical resources, we found physical infrastructure, like toilets, were adequate at health facilities, enabling pregnant women to provide urine samples. However, we found substantially lower availability of urine dipsticks among health posts compared to higher-level facilities in the NHFS analysis. Regarding competent, motivated human resources, we found that despite ANM’s knowledge on the importance of urine tests for conditions like detecting preeclampsia, few conducted urine tests at the point-of-care and few ordered urine tests at the recommended frequency according to the guidelines. Researchers often found the unavailability of lab assistants in health facilities due to various reasons, like leave, for training, resulted in pregnant women making repeated visits or being referred to higher-level facilities to obtain urine tests.

According to Nepal’s Basic Health Services Package, urine tests are part of basic health services and as per Nepal Health Infrastructure Development Standards 2017 (NHIDS 2017), urine tests are required to be accessible at a minimum of one health facility in each local government area(26,27). The selected facilities in our study also had urine tests available, except for one, indicating urine test availability in all three local government areas (municipalities). However, we found a substantial differences in the availability of urine protein and glucose tests from PHCC (more than 90%) to Health Posts (less than 30 %), highlighting inadequacy in readiness of lower-level health facilities to provide urine tests.

We identified gaps between guideline and practice in offering urine tests at the recommended frequency and in counselling about the tests’ purpose. Similar findings were found in a cross-sectional study evaluating ANC service provision in three hospitals in Nepal; many pregnant women reported receiving insufficient information about the purpose and procedures of various ANC services, including urine testing(28). The 2018 Right to Safe Motherhood and Reproductive Health Act states the right to receive appropriate counseling related to healthcare(30). However, interviews with pregnant women and researchers’ observations revealed a lack of appropriate communication between ANMs and pregnant women explaining the urine testing procedure and its results.

Nepal’s National Standard for WASH in Health Care Facilities mandates the WASH infrastructure in the form of functional toilets and hand wash basins, including soap as one of the necessary WASH items for infection prevention(28). This study also matched national guidelines, having functional toilets and wash basins in all health facilities, which is a facilitator for the conduct of urine tests too.

Though, urine dipstick tests method cannot be considered as a diagnostic method for detection of proteinuria as a marker of renal insufficiency or renal target organ damage, it remain a vital tool in resource-limited settings to screen preeclampsia and other complications and is performed by a range of health cadres (29,30,31,32). This study also found the conduct of urine dipstick test but relying on a lab assistant. We a reliance on lab technicians to perform urine dipstick tests and that few facilities had any ANC provider receiving training in ANC screening within the past 24 months. The lack of lab services availability and lack of ANC providers performing the test, particularly in lower level facilities that often serve rural women, create a major barrier for screening of preclampsia, gestational diabetes and urinary tract infection, resulting referrals to higher-level facilities and potential delays in early treatment.

### Strength & Limitation of the study

Our study offered several strengths in drawing from more in-depth qualitative findings in three municipalities in Kavrepalanchok District, Nepal, alongside nationally representative health facility survey data, but it was not without limitations. The small number of health facilities and interviews in the qualitative data, done in one district, hinders the generalizability of findings and comparability with the national data. Variations in practices across the different facilities made it challenging to develop conclusions, though we tried to mitigate this by sharing and refining our analysis in the July 2024 stakeholder workshop. The interviews conducted with healthcare providers and pregnant women were limited in scope due to time constraints of participants; the researchers encountered difficulties in encouraging participants to speak freely about their experiences and views. Further, the presence of researchers may have influenced behavior during observations in health facilities. Our study focused on urine testing and did not address other, potentially interrelated, aspects of the provision of antenatal care.

## Conclusion

Many pregnant women visit health posts and other grassroot level primary care centers (Community Health Units & Urban Health Centers) as their first contact point for routine check ups. Therefore, it is important to strengthen lab capacity at lower-level facilities or encourage ANC providers like ANMs to conduct the point-of-care testing for urine glucose and protein. Point of care urine testing could be an opportunity to align with guidelines ( ANC to PNC continuum of Care) and could meet the indicator of urine testing for protein in monthly ANC visits. Training and supportive supervision to ANMs for the point of care testing for urine glucose and protein and counselling on its purpose and outcome could strenghten quality ANC and support in screening, detection and management of non-communicable diseases like preeclampsia and gestational diabetes mellitus during pregnancy.

## Data Availability

Data supporting the findings of this study are available from the corresponding author on request

## Acknowledgement

We are deeply grateful to the municipalities health section chiefs, Antenatal care healthcare providers and pregnant women who participated in the interviews.

## Supporting information

**S1 Fig1: Data Sources included in the analysis mapped to the WHO framework for the quality of ANC in terms of urine testing. ANC =Antenatal Care, KII-Key Informant Interview, IDI-In Depth Interview**

**S2 Facility Observation Tool**

